# Exercise-mediated biomarker signatures from a combined aerobic and strength training intervention in Singaporean breast cancer patients: findings from the BREXINT Pilot Study

**DOI:** 10.64898/2026.08.17.26360564

**Authors:** Patrick Henry Sebastian Sitjar, Parthiban Periasamy, Si Ying Tan, Mabel Wong, Marek Kukumberg, Sabrina Adam, Joe Yeong Poh Sheng, Elaine Hsuen Lim, Jorming Goh

**Author notes:** Corresponding Authors: Elaine Hsuen Lim, National Cancer Centre Singapore (NCCS) Singapore 168583, Jorming Goh, Healthy Longevity Translational Research Program (HLTRP) Yong Loo Lin School of Medicine National University of Singapore (NUS) Singapore 117456.

## Abstract

Biomarkers perturbed by exercise-mediated molecular mechanisms, in women with early-stage (stage I-III, non-metastatic) breast cancer are poorly defined, and especially in under-represented Asian cohorts. In this exploratory Breast Cancer Exercise Intervention (BREXINT) pilot study, 15 Asian women were randomized to a combined aerobic and resistance exercise intervention program (n=8) and a control group (n=7). Fasting blood sampling was performed at baseline, 8,16, and 24-week timepoints. Blood parameters were imputed, transformed and screened for intervention-specific variations using IQR-trimmed, paired Wilcoxon tests. Twenty-one blood parameters were found to meet a differential change rule (significance observed in 1 group but not the other). Exercise-associated signatures displayed hematological and cytokine remodeling at 16-weeks. Control-associated signatures include adipokine and renal markers at 16 and 24-weeks. Of note, exercise-driven decrease of IL-10 at 16-weeks (*p*=0.022) retained significance following linear mixed effects confirmation among screened candidates. IL-10-centred modulation is the most convergent exercise-associated blood derived signature but warrants further validation in larger exercise oncology trials.

## Introduction

Breast cancer is the leading cancer among women in Singapore, accounting for almost a third of female cancer diagnoses, and ranks among the most commonly diagnosed cancers in women globally [1]. Advances in detection and treatment methods have improved patients’ survival rates but suffer from lower quality of life following the disease and treatment effects [2]. Women completing oncological therapy for early-stage (stage I-III) breast cancer frequently suffer from treatment-associated sarcopenia, cardiometabolic risk and low-grade systemic inflammation [3]. Currently, post-treatment follow-up remains weighted toward cancer surveillance and monitoring, while rehabilitation such as structured exercise training take a backseat [4]. Exercise training is widely advocated as supportive care for this population, with meta-analyses of randomized trials reporting that combined aerobic and resistance training can modulate pro-inflammatory mediators including IL-6 and TNF-α [5, 6]. Likely, exercise mitigates obesity, hormones and improves immune function to regulate cancer risk factors [7, 8].

Chronic inflammation is a key driver in cancer development and exercise training presents as a viable, non-pharmacological approach to regulate inflammation [9]. A pooled analysis by Meneses, *et al.* of 8 impactful exercise trials (n=478) found that following exercise interventions (8-24 weeks), no significant change was detected in circulating IL-10 and CRP but significant decrease in IL-6, TNFα, IL-8, and increase in IL-2[6]. These shed light on the possible inflammatory mechanisms that can be restrained by exercise training. IL-6 and TNFα are prominent biomarkers of interest in breast cancer studies due to its association with unfavorable prognoses in breast cancer [10]. IL-6 has been identified to trigger the JAK/STAT3 pathway which implicates breast cancer tumor initiation, growth, survival, metastases and further quell anti-tumor immune response while TNFα likely promotes epithelial-mesenchymal transition (EMT), metastasis and oncotherapy resistance [10–12]. However, such chronic exercise training studies in cancer patients are further complicated by variations in the exercise interventions from protocols, duration, frequency, modality and perhaps inter-individual response to exercise stimulus [6, 13, 14].

Most biomarker studies have enrolled predominantly Western cohorts, used conventional between-group comparisons at fixed timepoints, and under-report how missing longitudinal samples and multiplicity affect interference in small mechanistic trials [15, 16]. So far, biomarker responses in early-stage, non-metastatic Asian cohorts are scarcely reported, and effects on individual cytokines are heterogenous.

In this pilot study under Breast Cancer Exercise Intervention (BREXINT), cardiorespiratory fitness, body composition, and circulating immune and metabolic biomarkers were evaluated at serial timepoints during the intervention period, and compared between an intervention group and a control group [17]. Study participants were Asian women in Singapore with stage I-III breast cancer who had completed curative surgery, chemotherapy (if given) and radiotherapy (if given). The intervention comprised 24 weeks of supervised combined aerobic and strength training. In this article, we report an exploratory combined longitudinal approach with frequent sampling which showed specific exercise-mediated adaptations across blood parameters rather than a change observed in both groups. A data analysis pipeline was curated to integrate missing-data imputation, global visualization, differential screening with effect sizes, linear-mixed effects (LME) confirmation, imputation sensitivity and exercise-adherence dose-response evaluation. Further, this study probed both routine clinical parameters together with clinically validated inflammatory parameters such as neutrophil-to-lymphocyte ratio (NLR), platelet-to-lymphocyte ratio (PLR), systemic immune-inflammation index (SII) in a lesser studied population undergoing exercise intervention [18, 19].

We identified IL-10 as a biomarker of interest in the exercise intervention group. As a pilot study, statistical power for findings in this work means it should only be interpreted as hypothesis-generating, consistent with other-accrued lifestyle intervention trials in early-stage breast cancer.

## Results

### Study participants and baseline characteristics

The current pilot study initially recruited 17 Singaporean women who have completed their treatment for early-stage breast cancer, with 2 patients withdrawing from the study before baseline assessments could commence (Figure 1). Baseline characteristics are shown in Table 1. The average age of the pilot study patients were 57.71 ± 10.86 years, which was generally balanced between both the control and exercise intervention groups. Most of the study patients were ethnically Chinese (73.33%) and was diagnosed with Luminal A (ER+ and/or PR+, HER2-) breast cancer. All patients had curative breast surgery in combination with other therapies such as chemotherapy, radiotherapy, hormonal therapy, HER2 therapy and monoclonal antibody therapy. Of the 15 patients, 7 were randomized into the control group and 8 into the exercise intervention group where they took part in the structured exercise training intervention.

**Figure 1.**
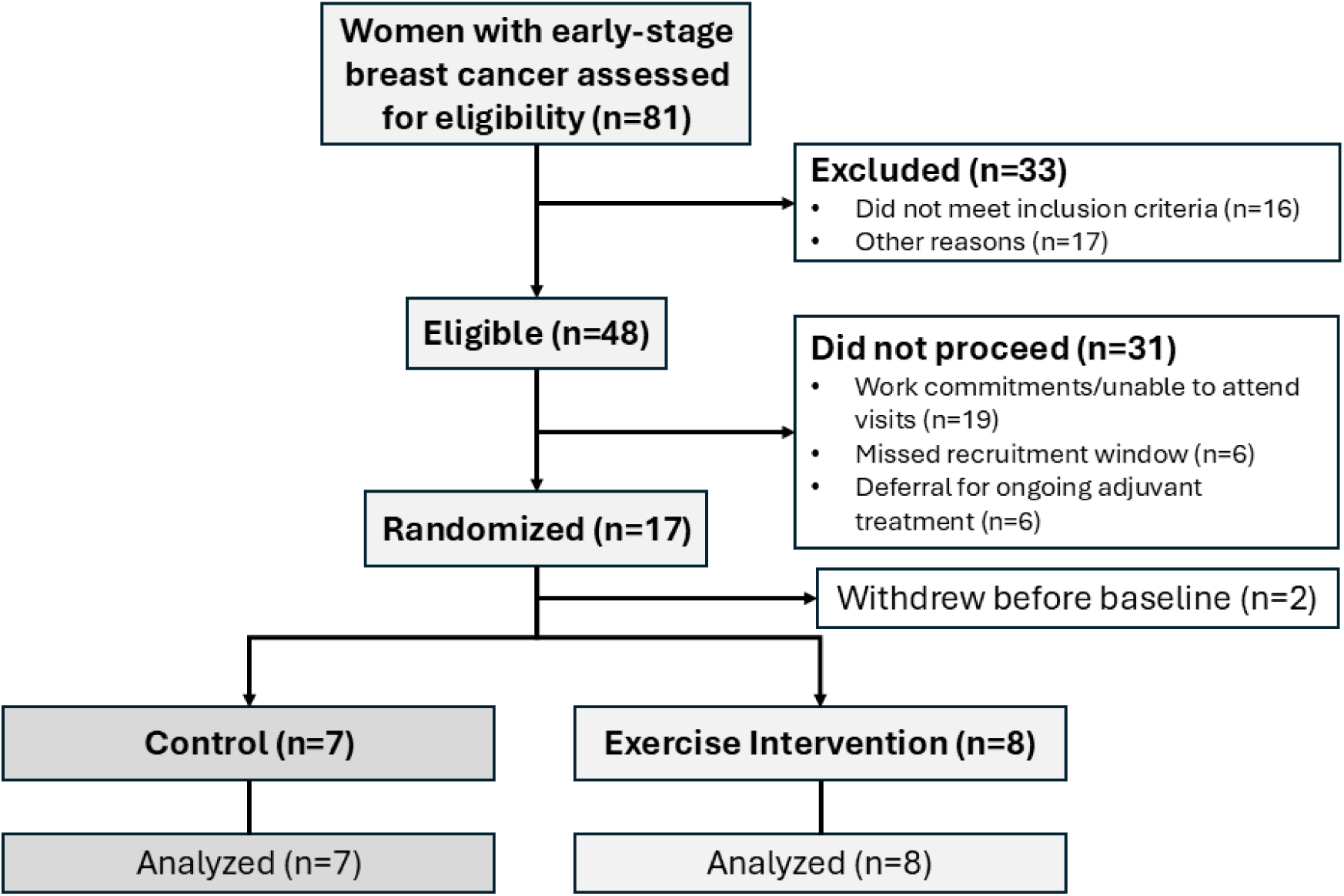
CONSORT Flow chart of the Singaporean breast cancer patients evaluated in the BREXINT Pilot Study.

**Table 1.** Baseline patient characteristics comparing the control and exercise intervention groups.

|  | All (n=15) | Control (n=7) | Exercise (n=8) |
| --- | --- | --- | --- |
| Age (years) | $57.71 \pm 10.86$ | $56.75 \pm 10.58$ | $57.65 \pm 11.27$ |
| Age range (years) | 42.83 – 73.32 | 47.26 – 68.22 | 42.83 – 73.32 |
| **Race, n(%)** |  |  |  |
| Chinese | 11 (73.33%) | 4 (57.14%) | 7 (87.50%) |
| Malay | 2 (13.33%) | 2 (28.57%) | 0 |
| Indian | 1 (6.67%) | 1 (14.29%) | 0 |
| Eurasian | 1 (6.67%) | 0 | 1 (12.50%) |
| **Breast cancer type, n(%)** |  |  |  |
| Triple Negative | 1 (6.67%) | 1 (14.29%) | 0 |
| Luminal A | 10 (66.67%) | 5 (71.43%) | 5 (62.50%) |
| Luminal B | 2 (13.33%) | 0 | 2 (25.00%) |
| HER2-Enriched | 2 (13.33%) | 1 (14.29%) | 1 (12.50%) |
| **Treatment history, n(%)** |  |  |  |
| Surgery | 15 (100.00%) | 7 (100.00%) | 8 (100.00%) |
| Chemotherapy | 14 (93.33%) | 6 (85.71%) | 8 (100.00%) |
| Radiotherapy | 10 (66.67%) | 4 (57.14%) | 6 (75.00%) |
| Hormonal therapy | 13 (86.67%) | 6 (85.71%) | 7 (87.50%) |
| HER2 therapy | 4 (26.67%) | 1 (14.29%) | 3 (37.50%) |
| Monoclonal antibody therapy | 4 (26.67%) | 1 (14.29%) | 3 (37.50%) |
| **Anthropometry** |  |  |  |
| Height (m) | 1.59 ± 0.06 | 1.59 ± 0.06 | 1.60 ± 0.06 |
| Weight (kg) | 56.65 ± 9.86 (n=14) | 57.37 ± 9.88 | 57.92 ± 9.01 (n=7) |
| Body mass index (kg/m<sup>2</sup>) | 22.11 ± 3.56 (n=14) | 22.50 ± 3.40 | 22.48 ± 3.43 (n=7) |
| **Menopausal status, n(%)** |  |  |  |
| Premenopausal | 7 (46.67%) | 3 (42.86%) | 4 (50.00%) |
| Postmenopausal | 8 (53.33%) | 4 (57.14%) | 4 (50.00%) |
| **Medical history, n(%)** |  |  |  |
| Smoking history | 1 (Ex-smoker) (6.67%) | 0 | 0 |
| Diabetes | 3 (20.00%) | 2 (28.57%) | 1 (12.50%) |
| Hypertension | 3 (20.00%) | 2 (28.57%) | 1 (12.50%) |
| Hyperlipidemia | 5 (33.33%) | 4 (57.14%) | 1 (12.50%) |
| Others | 1 CKD (6.67%)<br>1 Hypothyroidism (6.67%) | 1 Hypothyroidism (14.29%) | 1 CKD (12.50%) |

Data is presented as mean ± standard deviation with percentages in the parentheses representing % of the sample and group sizes respectively. Triple Negative (ER-, PR-, HER2-); Luminal A (ER+ and/or PR+, HER2-); Luminal B (ER+ and/or PR+, HER2+); HER2-Enriched (ER-, PR-, HER2+) ER: Estrogen Receptor PR: Progesterone Receptor HER2: Human Epidermal growth factor Receptor 2 CKD: Chronic Kidney Disease

### Global overview of blood, exercise and physiological parameters

The analysis matrix visualizes the longitudinal fluctuations in anthropometric parameters, exercise attendance (for the exercise intervention group), exercise assessments, summary of patient clinical history, and imputed, transformed blood parameters from the 15 BREXINT pilot study patients across baseline to 24-weeks and split by intervention group (Figure 2). Visually distinct row clusters spanning hematological indices, acute-phase proteins and cytokine/adipokine measures motivated formal intervention-specific screening.

**Figure 2.**
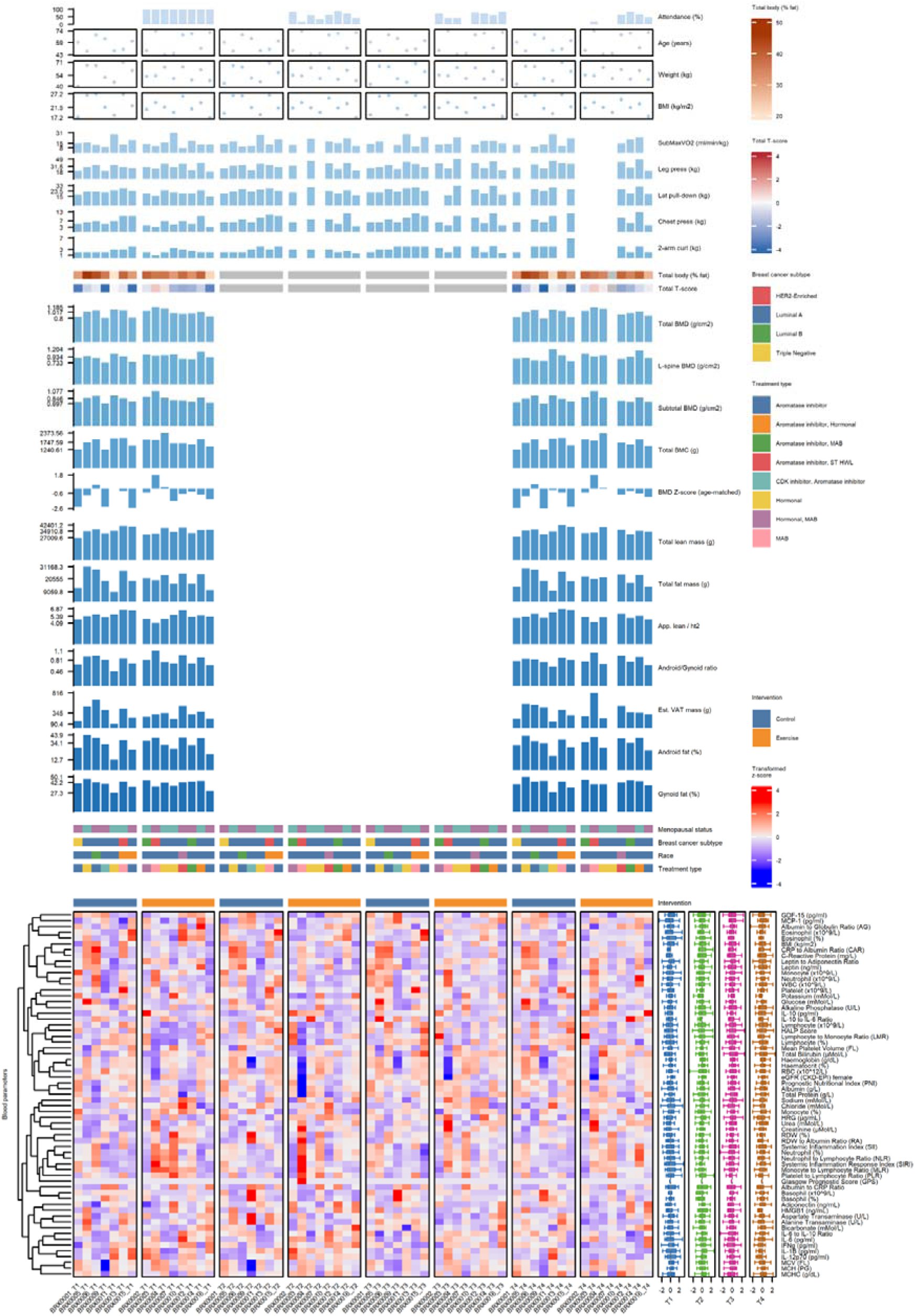
Global longitudinal overview of BREXINT patient data. Columns are ordered by timepoint (T1-T4) and split by intervention group (Control in Blue; Exercise in Orange). Complex heatmap indicates transformed z-score with boxplots summarizing each parameter by visit.

### Intervention-specific biomarkers of interest highlighting exercise-training adaptations

Comparison of paired Wilcoxon signed-rank analysis of parameters at baseline (T1) versus 8-weeks (T2), 16-weeks (T3) or 24-weeks (T4) between the control and exercise groups identified 21 blood-derived biomarkers of interest that showed statistically significant intervention-specific adaptations (Figure 3a & 3b).

**Figure 3.**
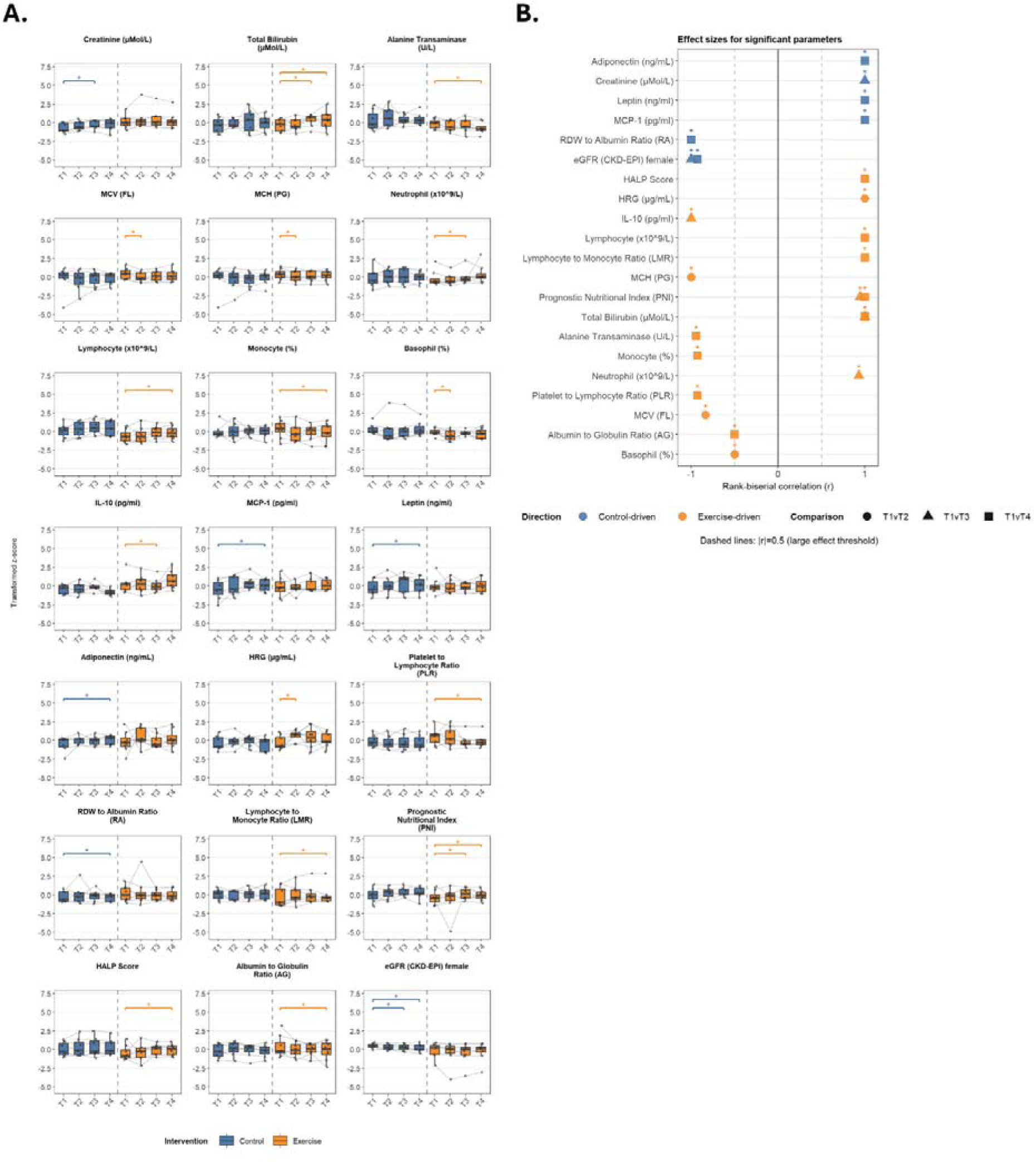
Differential biomarker changes in 21 screened parameters. A. Boxplots of transformed z-scores with the Control group on the left (in blue) and the Exercise intervention group on the right (in orange) with grey lines connecting individual participants. Significance brackets reflect paired Wilcoxon signed-rank tests on the intervention-specific side (*p*<0.05). B. Rank-biserial effect sizes (*r*) for significant comparisons with the Control group (in blue) or Exercise intervention group (in orange) with shape indicating T1 vs T2 (circle), T1 vs T3 (triangle) or T1 vs T4 (square). Dashed vertical lines denote |*r*|=0.5.

Exercise-induced liver and hematology parameter changes from T1 include elevated total bilirubin at T3 (*p*=0.022, r=1.00) and T4 (*p*=0.036, r=1.00), decrease in alanine transaminase at T4 (*p*=0.020, r=-0.944), lower MCV (*p*=0.042, r=-0.83) and MCH (*p*=0.022, r=-1.00) at T2. Exercise-driven variations in immune cell counts were also observed with elevated neutrophil counts at T3 (*p*=0.035, r=0.929) and lymphocyte counts at T4 (*p*=0.022, r=1.00), decrease in monocyte percentage at T4 (*p*=0.035, r=-0.929) and basophil percentage at T2 (*p*=0.036, r=-0.50). Derived inflammatory markers such as platelet-to-lymphocyte ratio (PLR) decreased at T4 (*p*=0.035, r=-0.929) and lymphocyte-to-monocyte ratio (LMR) increase at T4 (*p*=0.036, r=1.00) [18, 20]. Composite blood-derived prognostic biomarkers including the prognostic nutritional index (PNI) were elevated at T3 (*p*=0.021, r=0.944) and T4 (*p*=0.022, r=1.00), HALP score (hemoglobin, albumin, lymphocyte, platelet) was elevated at T4 (*p*=0.022, r=1.00) and albumin-to-globulin ratio was lower at T4 (*p*=0.036, r=-0.50) [21–23]. Finally, research biomarkers such as interleukin 10 (IL-10) decreased at T3 (*p*=0.022, r=– 1.00) and histidine-rich glycoprotein (HRG) was greater at T2 (*p*=0.014, r=1.00).

In comparison, certain parameters fluctuations were flagged specifically in the control group. Control-specific clinical and derived parameters include higher creatinine at T3 (*p*=0.022, r=1.00), RDW-to-albumin ratio (RA) decreased at T4 (*p*=0.036, r=-1.00) and estimated glomerular filtration rate (eGFR) (CKD-EPI) decreased at T3 (*p*=0.036, r=-1.00) and T4 (*p*=0.035, r=-0.929). In addition, research biomarkers including MCP-1 (*p*=0.036, r=1.00), leptin (*p*=0.036, r=1.00), and adiponectin (*p*=0.036, r=1.00) were all increased at T4.

### Robustness analyses nominate IL-10 as the leading exercise-associated signal

To further evaluate the 15 exercise-specific parameters, delta from baseline (z-score) plotted against the exercise attendance (%) at the comparison timepoint showed conflicting observations with HRG at T2 displaying a supportive attendance trend (*p*=0.086, Spearman rho=0.64) but lymphocyte count at T4 showing a significant negative correlation (*p*=0.049, Spearman rho=-0.76) (Figure 4a.). Among the selected parameters, a change-score correlation network using Spearman rho ≥0.5 for each intervention group show which parameters are simultaneously perturbed (Figure 4b.). To ensure the robustness of the identified parameters, the imputation sensitivity was plotted to rule out parameters that depended on imputed datapoints (Figure 4c.). 13 of the 21 parameters were retained including IL-10, creatinine, MCP-1, leptin, adiponectin and eGFR but 8 were not, namely HRG, PLR, LMR, monocyte percentage, HALP score, lymphocyte count, alanine transaminase and albumin-to-globulin ratio. Finally, LME model confirmation determined that only IL-10 had composite robustness as an exercise-associated signal (*p*=0.029) while other candidates such as HRG and lymphocyte count might be associated with attendance of training sessions, but should be interpreted with caution, *p*>0.05.

**Figure 4.**
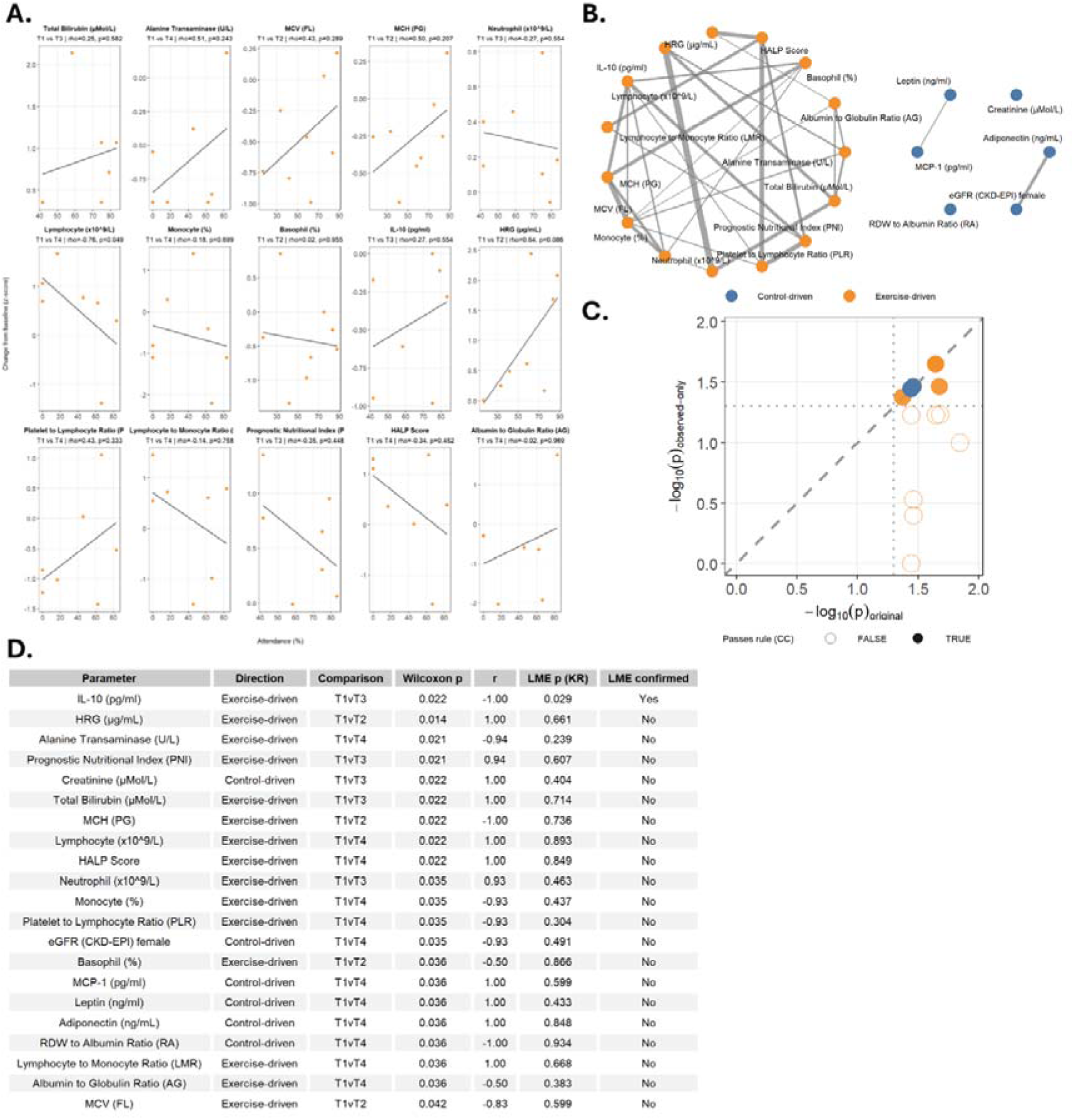
Robustness of 21 screened biomarker signals. A. Scatterplot representing 15 exercise intervention group specific parameters (delta from baseline, z-score) against exercise attendance (%). B. Change-score correlation network among screened parameters with edges drawn when Spearman rho ≥ 0.5 within the same intervention group. C. Imputation sensitivity assessment comparing *log*10(Original Wilcoxon P) vs –*log*10(observed-only P) after masking imputed data points. D. Linear mixed-effects (LME) model confirmation for 21 biomarker candidates assessed by Group x Time interaction models (Kenward-Roger P; confirmation flag).

## Discussion

In this exploratory pilot study in an Asian, Singaporean cohort of breast cancer patients following cancer treatment, we identified that a combined exercise intervention prompted fluctuations in several hematological, immune and inflammatory biomarkers. It is well characterized that following debilitating cancer therapy, patients are prone to cancer-related fatigue (CRF) and some degree of skeletal muscle degeneration [24]. In addition, while common cancer treatments like surgery, chemotherapy and radiotherapy can successfully treat or control disease progression, they are not without detriment. These treatments insults healthy and cancer cells alike, causing oxidative stress and an upregulation of inflammatory proteins leading to musculoskeletal weakness which can be further compounded by extended bed rest following treatment [25–27]. Toxic cancer therapies which are known to perturb the HPA axis and endocrine system, and introduce mitochondrial abnormalities which further drive inflammation [28].

Exercise training exerts a potent pleiotropic, anti-inflammatory effect in the tumor microenvironment (TME) and may further prime immune cells against tumor cells [7, 29]. Also, exercise training has shown its efficacy in alleviating CRF and maintaining muscle mass [30, 31]. These anti-cancer benefits can be attributed to exercise-induced molecules or ‘exerkines’ that have been proposed to directly act on cancer cells or by altering the TME. Literature review of published studies in exercise oncology have identified cytokines and immune markers such as IL-6, irisin, SPARC, oncostatin M (OSM), decorin and even changes in immune cell subsets but these findings are usually heterogenous [29, 30]. Likely, acute exercise triggers an increase in the above-mentioned cytokines that drive upregulation in immune cells such as CD4, CD8 T cells, NK cells, B cells and monocytes [7, 32]. Over time, exercise-induced immune markers and receptor homeostasis shifts, which are highly varied in different individuals which makes it difficult to identify biomarkers for cancer patients in exercise trials. Based on longitudinal analyses of blood markers in both exercise intervention and control groups across 24 weeks, we have also observed a greater variation in hematological, immune and inflammatory markers in the exercise intervention group while there are clinical variations specific to the control group.

Of note, IL-10 was screened as a biomarker of interest with support from differential Wilcoxon screening, masking of imputed values and a significant LME model confirmation. As such, could serve as a key biomarker in exercise intervention studies for cancer populations. IL-10 is a pleiotropic anti-inflammatory cytokine that restrains immune cell activation and inhibits the production of pro-inflammatory cytokines such as IL-6 and TNFα [33, 34]. Mechanistic studies suggest that IL-10 regulates inflammation duration through sphingolipid metabolism whereby fatty acid desaturation occurs and prevents very-long-chain (VLC) ceramides accumulation which drives macrophage inflammation [34]. Its circulating concentration after chronic exercise is inconsistent across oncology trials [6, 35–37], making directionality biologically informative rather than self-evident. The exercise intervention group showed a decrease at T3, which suggests a recalibration of the immune homeostasis. For example, regular muscle contractions during exercise reduces the necessity for compensatory anti-inflammatory response as pro-inflammatory monocyte programming decreases [38, 39]. Further functional immune assays are needed to distinguish these mechanisms.

HRG and other hematological indices increased at T2 that could suggest broader systemic adaptations at play. MCP-1, leptin and adiponectin increased specifically in the control group may indicate natural history, treatment effects or possible that exercise training could partially buffer adipose-inflammatory cross talk during early recovery. This pattern contrasts with acute exercise studies reporting that transient MCP-1 elevation post-session in healthy women [11] and is more consistent with CCL2/CCR2 chemokine biology in cancer [40].

Exercise-driven shifts in PNI, HALP score, PLR and LMR are also consistent with hematological and nutritional recalibration after chemotherapy [6], but these composite indices did not pass LME confirmation in our analysis so it may likely reflect our low sample size screening sensitivity rather than independent confirmatory signals.

While there are directional disparities in the identified immune markers with other reported studies, these could be attributed to homeostatic adaptations to compounding changes brought about by the initial breast cancer disease, its treatments and for the exercise-intervention group, the additional inflammatory stress conferred by exercise training [41]. Further complicated by varying adherence to the exercise intervention program and highly individualized molecular response to exercise training, pinpointing directionality in exercise-induced biomarkers presents a significant challenge [42].

There is a growing base for personalized exercise prescription, especially for cancer patients who have highly heterogenous diseases and physical makeup. IL-10 concentrations could serve as a valuable biomarker to monitor breast cancer patients’ immune homeostasis during recovery following treatment and as they navigate exercise training. Further work is needed to validate this in larger scale breast cancer exercise cohorts.

This study has important limitations. Recruitment only reached 15 participants, which limits statistical power. The main challenge identified from recruitment was that following cancer treatment completion, patients needed to return to work as well as obtain medical exemption to participate in group exercise interventions. This emphasizes that Asian women have a younger age of onset with breast cancer [43]. As the pilot study mandated in-person supervision of the exercise sessions, which took place during office hours, this immediately excluded those in the workforce. Exercise adherence was also heterogeneous and relatively poor due to patients’ commitments elsewhere. Further work should be flexible to meet the demands of cancer patients following treatment and as they navigate post-cancer life. Further, the cohort was recruited from a single center and predominantly Luminal A type BC.

Although blood parameters and mainly IL-10 immune calibration were identified in the exercise group and adipokine, chemokine and renal drifts were found in the usual care group. Further work can investigate functional immune phenotyping to determine exercise specific adaptations.

These findings should therefore be interpreted as hypothesis-generating and require further validation in larger, adequately powered cohorts. Once validated, exercise-induced IL-10 immune modulation dynamics could present a reproducible signature of structured exercise in non-metastatic breast cancer rehabilitation.

## Methods

### Study design and participants

The BREXINT study is a prospective, randomized pilot clinical trial studying the effects of a 24-week combined aerobic and strength training program against usual care in women with stage I-III (early-stage, non-metastatic) who have completed curative treatment regimens. The study protocol was published previously. Briefly, these women were aged ≥21 years and enrolled within 8 weeks of their final treatment if they met the inclusion criteria and excluded if they had medical issues that might be unsuitable for exercise training as determined by the investigators. The pilot study patients were randomized into either the exercise intervention group which participated in aerobic and resistance training sessions (up to 3 sessions per week) for 24 weeks at the Singapore Cancer Society (SCS) rehabilitation gym, overseen by physiotherapists and cancer exercise trainers while the control or usual care group were only provided details about the training program. Study patients were subjected to blood sample collection, exercise testing and questionnaires at baseline (T1), 8 (T2), 16 (T3) and 24 (T4) week timepoints with body composition assessed with dual x-ray absorptiometry (DEXA) at T1 and T4 only.

### Ethics Approval

All procedures and assays were performed in accordance with the Declaration of Helsinki. The BREXINT trial was registered at ClinicalTrials.gov (NCT05957068) and was approved by the following institutional review boards (IRBs): the National University of Singapore (NUS) IRB (NUS-IRB-2022-700), SingHealth Centralized Institutional Review Board (CIRB) (2020-2451), and the Agency for Science, Technology and Research (A*STAR) IRB (2022-079). Written consent was obtained from all participants before commencing data collection.

### Exercise assessments

All exercise assessments were conducted at the SCS rehabilitation gym. Submaximal VO_2peak_ following a modified Balke protocol on a treadmill was carried out to determine cardiorespiratory fitness. The patients were connected to a metabolic cart (Metalyzer 3B, Cortex Biophysik, Germany) to collect physiological parameters while undergoing a graded exercise test on the treadmill. Briefly, the test comprised of a 5 min rest to collect baseline readings before proceeding onto the treadmill for a warm-up at 3.2 Km/hr at 0% incline for 1 min, followed by 4.3 Km/hr at 0% incline for 1 min and a final minute at 4.8 Km/hr at 1% incline. Following the warm-up, the speed was kept at 4.8 Km/hr but the incline increased by 1% every 1 min until the patient did not want to proceed or once 85% of age-predicted maximal heart rate (MHR) (220-age in years) was reached.

Muscle strength was assessed by determining the 10-repetition maximum (10RM) for chest press, latissimus pull down, 2 arm curl and leg press.

Blood pressure was monitored before and after the exercise assessment sessions for patient safety in accordance with the American College of Sports Medicine (ACSM) guidelines for exercise testing in clinical populations.

### Exercise training

The exercise intervention group was scheduled to carry out scheduled exercise sessions on Mondays, Wednesdays and Fridays which consisted of 20 min treadmill walking with participants maintaining a heart rate of between 50-75% MHR. Resistance training comprised of chest press, bicep curls, shoulder flexion and leg press on Mondays; dumbbell rows, elbow extension, upright row and sit to stand on Wednesdays; squats, bridging, hip extension and hip abduction on Fridays. These exercises targeted the major muscle groups and progressed from 3-4 sets of 8-10 repetitions.

### Body composition

Body compositions were analyzed by DEXA (Hologic A, Hologic, Marlborough, Massachusetts, USA) in the Singapore General Hospital (SGH).

### Blood sample collection and laboratory assays

Fasting antecubital blood was collected at NCCS. Serum and whole blood samples were analyzed in SGH for routine clinical biochemistry and hematology tests. A separate whole blood sample was dispatched to the NUS laboratory and processed within 6 hr. Following centrifugation, plasma and buffy coats were aliquoted and stored in –80°C. Plasma immune and inflammatory biomarkers (IL-1β, IL-6, IL-10, IL-12p70, IFNy, GDF-15, MCP-1 and leptin) were quantified by Luminex multiplex immunoassay (LX200, Thermo Fisher Scientific, Waltham, Massachusetts, USA) with analyte concentrations extrapolated using a 5-parameter logistic (5PL) regression model. Commercial ELISA assays were performed to determine high mobility group box 1 (HMGB1) (Arigo ARG81185, Zhubei City, China), HRG (Cloud-Clone Corp. SEC534Hu, Houston, Texas, USA) and adiponectin (R&D Systems DRP300, Minneapolis, Minnesota, USA) concentrations. Buffy coat samples underwent DNA extraction (QIAGEN FlexiGene DNA kit, Venlo, Netherlands), followed by bisulphite conversion (ZymoResearch EZ DNA Methylation kit, Irvine, California, USA) and subsequently hybridized onto Infinium Methylation EPIC v2.0 array (Illumina Inc., San Diego, California, USA) and scanned using an iScan device (Illumina Inc., San Diego, California, USA). All commercial products were used following the protocols supplied by the manufacturers.

### Biomarkers derived from clinical blood results

Missing values from blood derived tests were first filled by within-subject linear interpolation across timepoints, then by block-wise MICE (m=5 imputations, maximum 10 iterations, seed 2026?). Additional inflammatory biomarkers including neutrophil-to-lymphocyte ratio (NLR), platelet-to-lymphocyte ratio (PLR), Systemic inflammation index (SII), RDW-to-albumin ratio (RA), lymphocyte-to-monocyte ratio (LMR), MLR, systemic inflammation response index, CRP-to-albumin ratio, PNI, HALP score, Glasgow prognostic score, leptin-to-adiponectin ratio, IL-6/IL-10 ratios, albumin-to-globulin ratio, BMI and CKD-EPI eGFR were calculated and analyzed. Each parameter was then independently transformed: z-score if skewness ≤1; log1p-z if skewed non-negatively; Yeo-Johnson-z if skewed with negative values.

### Statistics

Analyses were performed in R (v4.3) using BREXINT_Analysis_Script.R. Parameters were compared using paired Wilcoxon signed-rank tests at T1 vs T2/T3/T4 independently by intervention group on transformed z-score values following interquartile range (IQR) x 1.5 trimming (at least 4 retained values per intervention). Rank-biserial correlation r=1-4V/[n(n+1)]. A parameter passed screening if exactly one intervention group had *p*<0.05 and the other group had *p*>0.05 by comparison; no multiplicity adjustment was applied. Linear mixed-effects models (value ∼ Intervention x Timepoint + (1| StudyID); restricted maximum likelihood with Kenward-Roger *p* values) were fitted per screened parameter. Robustness analyses included Spearman correlation of exercise-driven change scores based on exercise attendance (%), within-intervention group change-score networks (|rho|≥0.5) and parameters were rescreened after masking imputed values. This exploratory pilot analysis used all available longitudinal blood data without prospective power calculation for parameter screening.

## Data and code availability

Data analysis code will be made available on Github (https://github.com/PatrickHenrySGH/BREXINT-Pilot-Study). Data and code will be made available upon request and subject to ethics restrictions. Access may be granted upon reasonable request with institutional approval.

## Acknowledgements

We would like to extend our sincere thanks to the Singapore Cancer Society staff; National Cancer Centre Singapore clinical research coordinators; National University of Singapore Exercise Physiology & Biomarkers Laboratory staff and students and the patients who participated in this pilot study. This work was supported by grants from the NUS Yong Loo Lin School of Medicine, Healthy Longevity Translational Research Program (HLTRP/2022/PS-03), National Cancer Centre Research Fund (NCCRF-YR2019-JUL-PG4). The funders had no role in study design, data collection, analysis, interpretation or manuscript preparation.

## Author contributions

M.W., E.H.S., and J.G. conceived and designed the research study; S.Y.T. and E.H.L. recruited patients; P.H.S.S., S.A. and M.K. performed testing and experiments; P.H.S.S., P.P., S.A. and M.K. analyzed data; P.H.S.S., P.P., M.K., S.A. and J.Y.P.S. carried out data interpretation; P.H.S.S. and P.P. prepared figures; P.H.S.S. and P.P. drafted manuscript; all authors reviewed and revised the manuscript; all authors approved the final draft of the manuscript.

